# Urine Proteome of COVID-19 Patients

**DOI:** 10.1101/2020.05.02.20088666

**Authors:** Yanchang Li, Yihao Wang, Huiying Liu, Wei Sun, Baoqing Ding, Yinghua Zhao, Peiru Chen, Li Zhu, Zhaodi Li, Naikang Li, Lei Chang, Hengliang Wang, Changqing Bai, Ping Xu

## Abstract

The atypical pneumonia (COVID-19) caused by SARS-CoV-2 is an ongoing pandemic and a serious threat to global public health. The COVID-19 patients with severe symptoms account for a majority of mortality of this disease. However, early detection and effective prediction of patients with mild to severe symptoms remains challenging. In this study, we performed proteomic profiling of urine samples from 32 healthy control individuals and 6 COVID-19 positive patients (3 mild and 3 severe). We found that urine proteome samples from the mild and severe COVID-19 patients with comorbidities can be clearly differentiated from healthy proteome samples based on the clustering analysis. Multiple pathways have been compromised after the COVID-19 infection, including the dysregulation of immune response, complement activation, platelet degranulation, lipoprotein metabolic process and response to hypoxia. We further validated our finding by directly comparing the same patients’ urine proteome after recovery. This study demonstrates the COVID-19 pathophysiology related molecular alterations could be detected in the urine and the potential application of urinary proteome in auxiliary diagnosis, severity determination and therapy development of COVID-19.

## INTRODUCTION

Coronavirus Disease-2019 (COVID-19) is caused by a novel virus strain, the severe acute respiratory syndrome coronavirus 2 (SARS-CoV-2), which is an unprecedented global health threat [1, 2]. Within just four months, a total of 2.47 million confirmed cases and nearly 170,000 fatalities, spreading in almost all countries and regions of the world, has been reported. Even worse, more than 70,000 new cases are being confirmed daily. However, no clinical drugs or vaccine is available for highly infectious SARS-CoV-2, which further exacerbates the panic.

Tremendous efforts have been devoted to investigating the SARS-CoV-2, and its host response, epidemiological and clinical characteristics to mitigate the current pandemic [1, 3–10]. Compared to the SARS-CoV spike (S) protein, the homologous SARS-CoV-2 S protein showed a higher affinity interaction with angiotensin-converting enzyme 2 (ACE2), which is thought to be the major receptor mediating the entry of the virus into the cell [11]. The SARS-CoV-2 has been reported to be harmful to lung, liver, heart, testis, bladder and kidney, where ACE2 are highly expressed [5, 8, 9, 12, 13]. Phylogenetic network analysis showed that the circulating SARS-CoV-2 is consisted of 3 types (A, B and C) and the origins and transition chains of the virus should be further validated [14]. It has been estimated that about 80% of COVID-19 patients experiencing mild symptoms (M-COVID), recover with, or even without conventional medical treatment [10]. However, the remaining 20% of patients with respiratory distress symptom may die rapidly without urgent and specialized intensive medical care, including immediate oxygen therapy, and mechanical ventilation [15, 16]. Disease stage significantly affects COVID-19 treatment and survivorship. The overall mortality rate for hospitalized patients varied from 2.3% for patients diagnosed at the early stage to 11% at the advanced stage [17]. Unfortunately, the majority of cases are diagnosed at the advanced stage due to the lack of biomarkers and medical resources at the early stage. Therefore, it is critical to develop novel approaches to estimate the disease stage for patients in order to seek appropriate treatments and allocate scarce medical resources. In addition, novel detection methods that genuinely reflect the underlying changes of molecular and biological processes of COVID-19 patients would be favorable to the understanding of SARS-CoV-2 pathogenesis.

Blood and urine are frequent biometrics for discovery of biomarkers of human diseases because of their accessibility and non-invasiveness. The compositions of proteins detected in blood and urine samples can genuinely reflect the changes of the body health conditions; thus, they are considered an important source for early warning and sensitive for disease detection [18, 19]. Recently, MS-based serum proteomics studies has been utilized to predict the severity of COVID-19 infection [20]. However, no study has been reported on the more accessible urine sample to date. Therefore, a comprehensive profiling of the urine proteome of COVID-19 patients will likely provide better diagnostics and clinical investigations of this disease.

In this study, we evaluated the diagnostic roles of urine samples on the progression of mild to severe type of COVID-19, and recovery state with cutting-edge urine proteomics [21]. Six COVID-19 patients, comprised of 3 diagnosed as severe cases including one death and 3 mild patients, were investigated. To confirm the findings derived from the urine proteome, two recovery samples were further analyzed. We found that proteins related with complement activation and hypoxia were highly up-regulated, while proteins associated with platelet degranulation, and glucose and lipid metabolic process were especially down-regulated in the COVID-19 severe type patients. However, the changed proteins during the infectious phase recovered to normal in the recovery stage. We propose that urine proteome characterization can be potentially used to distinguish and predict the COVID-19 progression of the mild to severe type. These urine proteome characteristics and changes may also shed light on the understanding of the COVID-19 pathogenesis.

## RESULTS

### Characterization of urine proteomes in controls and six COVID-19 patients

In total, we assayed 40 urine specimens that passed quality check (QC), including 32 healthy controls, 6 COVID-19 patients and 2 corresponding recovery person (Figures 1 and S1). All patients were tested positive for the presence of SARS-CoV-2 nucleic acid. They all developed either fever or cough. Severe patients showed typical symptoms of fatigue and dyspnea (Figure 1A). All patients had comorbidities, including 4 patients with essential hypertension, 1 patient with both essential hypertension and diabetes, and 1 patient with multiple metastases of colon cancer (dead on March 3, 2020) (Figure 1A). According to the Diagnosis standards [10], these six patients were categorized into two disease types: three patients were defined as severe type acute respiratory syndrome (S-COVID) and the other three were diagnosed as mild type (M-COVID).

**Figure 1.**
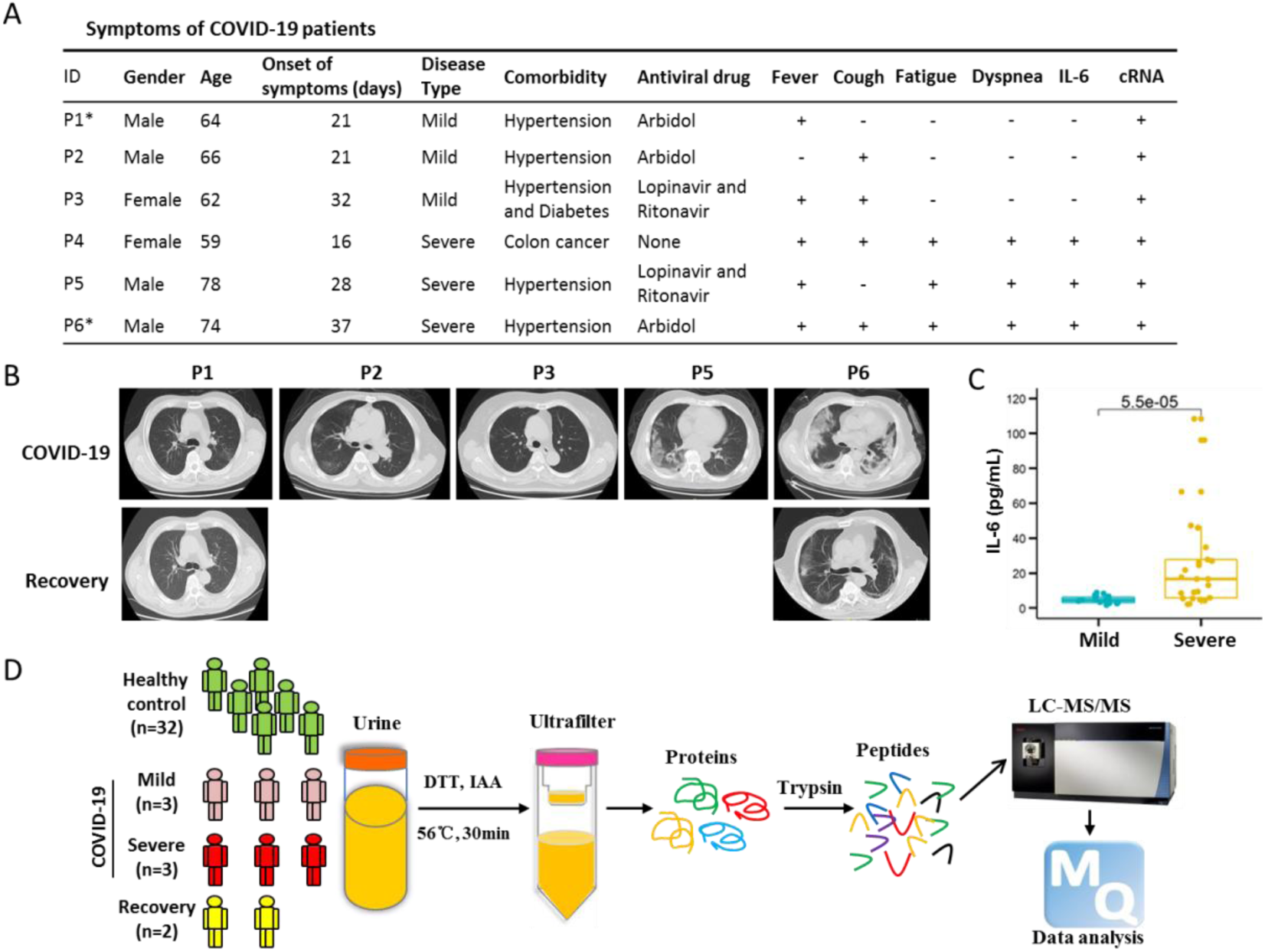
Proteomics study on urine samples of COVID-19 patients. (A) Basic information and clinical symptoms of COVID-19 patients, including mild (n=3) and severe (n=3) patients. No.4 patient (P4) was with multiple metastases of colon cancer and died on March 3, 2020. No.1 (P1) and 6 (P6) patients labeled with asterisk indicated the persons providing the recovery urine samples. cRNA indicated that the SARS-CoV-2 nucleic acid. (B) Ground-glass opacity on Computed Tomography (CT) of COVID-19 patients. (C) The amounts of IL-6 between mild and severe COVID-19 patients. (D) Experimental design of urine proteomics for COVID-19 patients. Interleukin-6, IL-6.

The severe COVID-19 patients showed ground-glass opacity in the lungs on Computed Tomography (CT) scanning (Figure 1B). After treatment, the lung shadow disappeared and gradually recovered (Figure 1B). Because the patient 4 (P4) had multiple metastases of colon cancer, only X-ray test was obtained (Figure S2). Interleukin-6 (IL-6) is an indicator of inflammatory storms (REF) [22]. We found the level of IL-6 in mild patients was 4.73 ± 2.03 pg/mL (mean ± standard deviation), while the expression level of IL-6 in severe patients was significantly higher than the normal standard (≤ 7.0 pg/mL) and drastically fluctuated during the infection, indicating that the stress response to viral infection in S-COVID patients was more severe (Figures 1C and S3).

The urine samples were collected after the diagnosis of the COVID-19. Four urine samples (H01-H04) of healthy controls were processed in parallel with the samples of COVID-19 patients (Figure 1D), which were further compared with the other healthy sample datasets (H05-H32) generated in the laboratory following the same sample preparation processes and mass spectrometry analysis in order to detect sample heterogeneity. To confirm the proteome shift observed from the COVID-19 patients, we also collected urine samples from two recovered patients (P1 and P6) (Figures 1A and S3).

As the sample size increases, the number of identified proteins in control group grows quickly, and gradually become saturated (Figure 2A). The peptide over protein ratio was 6.0 (Table S1), indicating high quality and reliability of our protein identification. To improve the accuracy of COVID-19 and recovery samples, 2 technical repeats were measured for each sample. A total number of 2656 proteins was identified from 32 healthy control samples (Figures 2A and Table S1). We identified and quantified 1380 and 1641 proteins in urine samples from COVID-19 and two recovery person in total, which was significantly lower than that of healthy controls (Figure 2B and 2C, Tables S2 and S3).

**Figure 2.**
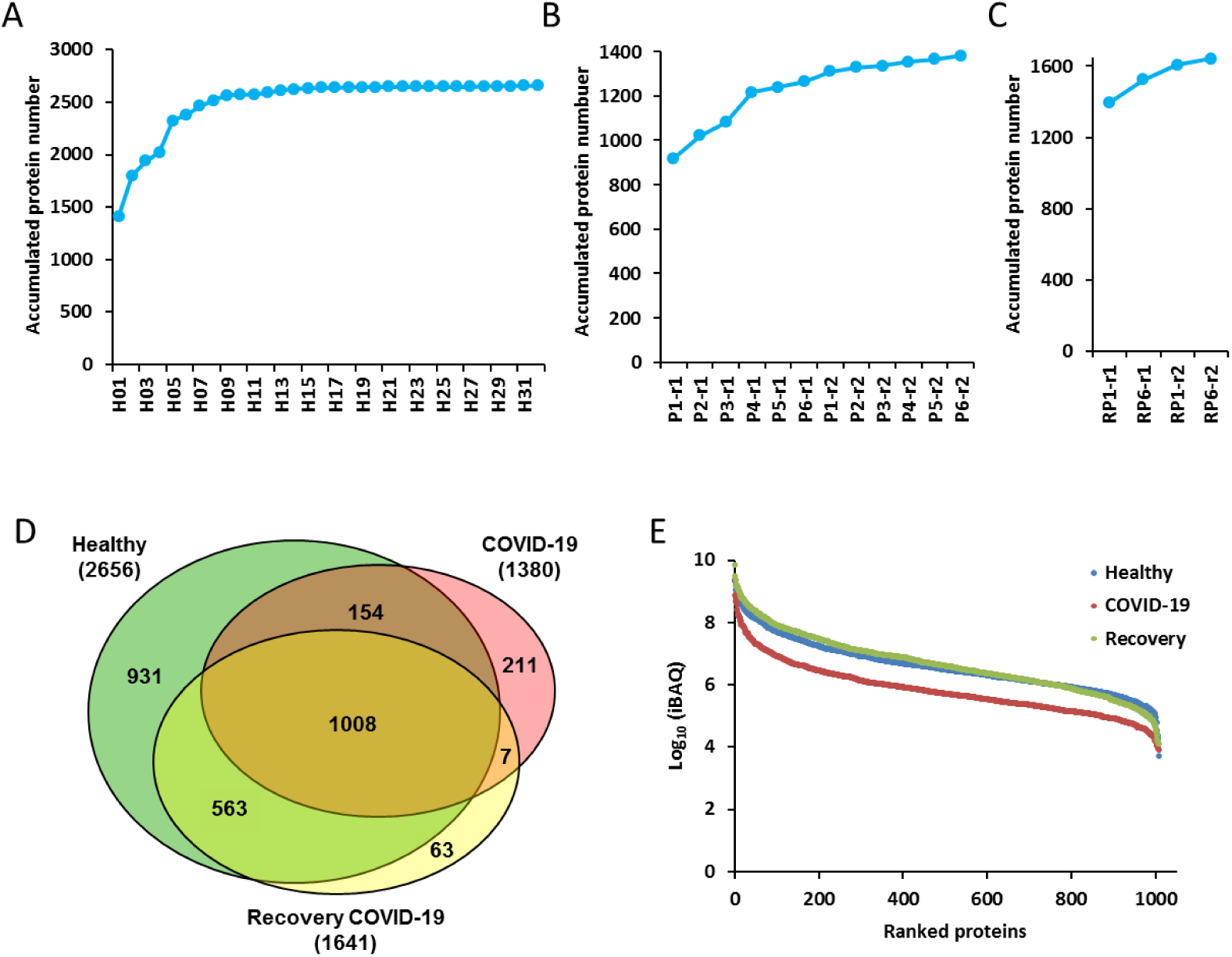
Identification and quantification of urine samples from COVID-19 patients and healthy controls. (A&B&C) The accumulation curve of the quantified proteins from 32 healthy volunteers (A), 6 COVID-19 patients (B) and 2 recovery patients (C). (D) The Venn diagram for the identified urine proteins from the healthy volunteers, COVID-19 and recovery patients. (E) The dynamic range of the iBAQ abundance of identified proteins from healthy volunteers, COVID-19 patients and recovery ones. The average abundance for each group was calculated.

There were 1008 proteins being commonly identified and quantified among the healthy controls, COVID-19 patients and recovered patients. However, 211 and 63 proteins were uniquely expressed in COVID-19 patients and recovery samples, respectively (Figure 2D). The average abundance of the identified proteins for each group spanned about 6 orders of magnitude, with lower abundance for the COVID-19 samples compared with healthy and recovery ones (Figure 2E). To check whether the SARS-CoV-2 proteins were present in the urine sample, we added SARS-CoV-2 protein sequences to the human proteome database, and no related proteins were identified. Low abundances of SARS-CoV-2 or its fragments in the urine or the relatively normal renal filtration function of the patients might explain the absence of SARS-CoV-2 protein in the urine samples.

### Urine proteomics differentiates COVID-19 patients from healthy people

To assess the quantitative variation and accuracy of the MS datasets, each urine sample of COVID-19 patients and the respective recovered samples were technically repeated twice. The absolute quantitative information iBAQ value was used for further comparison and analysis. The correlation coefficiency (R^2^) of the two replicates for each sample was higher than 0.80 (Figure S4), indicating the MS data were acquired with high degree of consistency and reproducibility in this study.

Due to the differences in sample size and operation during the sample processing, we found significant quantitative variations among different samples (Figure S5A). Therefore, the median values of iBAQ for each sample dataset were normalized equally to reduce the potential biases before quantitatively comparing the samples under COVID-19 with healthy conditions (Figure S5B). We also used the ComBat [23] to adjust for batch effects in datasets where the batch covariate was known. We found that the correlation of samples within the healthy and recovery groups were higher than that between the healthy and patient groups (Figure S6).

We found that patients and healthy people can be divided into two categories based on our cluster analysis (Figure 3), indicating the distinctive molecular characteristics between healthy and COVID-19 conditions. Interestingly, the urine samples of two recovery patients were clustered with healthy people (Figure 3). We also found that normal control individual H5 and H6 were “incorrectly” clustered with the M-COVID patients. Nevertheless, the M-COVID and S-COVID samples were clustered into separated groups except for one M-COVID patient (P3) with both hypertension and diabetes. The differences between M-COVID and S-COVID urine proteome samples might reflect different physiological responses of the COVID-19 infection at the proteome level.

**Figure 3.**
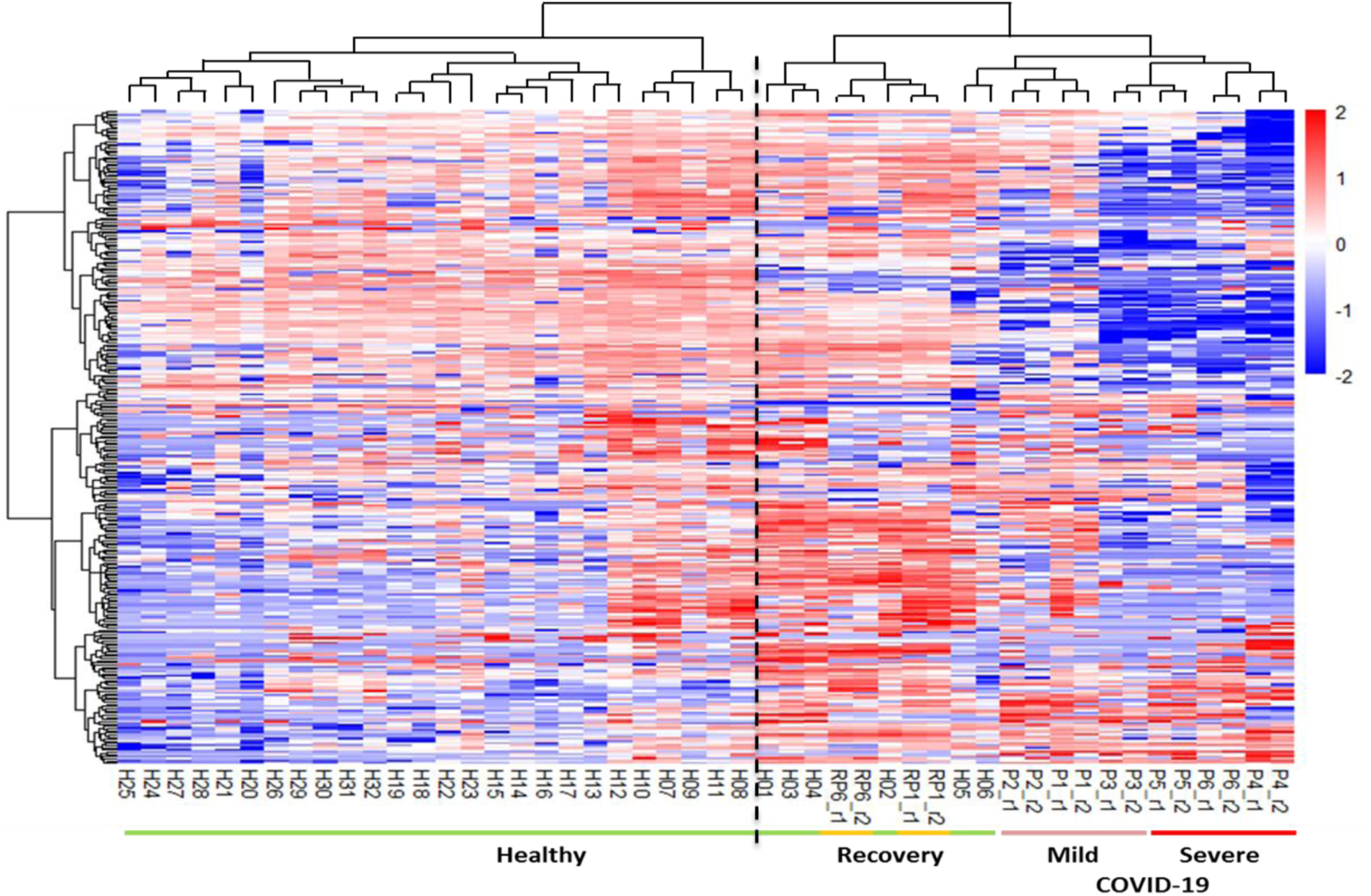
Distinction of healthy volunteers, COVID-19 patients and recovery patients in proteomic features. The clustering heatmap analyses differentiates healthy volunteers from COVID-19 patients and recovery ones.

### Molecular features of urine proteome for the pathogenesis of severe COVID-19 patients

Despite only 6 COVID-19 urine samples were tested, our results showed a clear distinction between healthy control and COVID-19 patient urine proteomes. We used the fold change ≥2 and *p*-value < 0.05 as filters to find the differentially expressed proteins between mild and severe diseases compared to the healthy control, respectively. There were 86 and 83 significantly up-regulated proteins, and 100 and 172 significantly down-regulated proteins in mild and severe COVID-19 samples, respectively (Figure 4A and 4B, Tables 4 and 5). To eliminate the confounding effects of the individual characteristics such as age or comorbidities, we also compared recovery samples with healthy controls and identified 278 differentially expressed proteins, including 152 up-regulated and 126 down-regulated proteins (Figure 4C and Table S6), which were excluded in the further analysis. We identified 95 unique changed proteins for severe type disease, 44 for mild type and 75 overlapped ones for both types of COVID-19 (Figure 4D). GO analysis of these changed urinary molecular features implied that the COVID-19 could result in the dysregulation of immune response, viral process, response to hypoxia, complement activation and platelet degranulation (Figure 4E).

**Figure 4.**
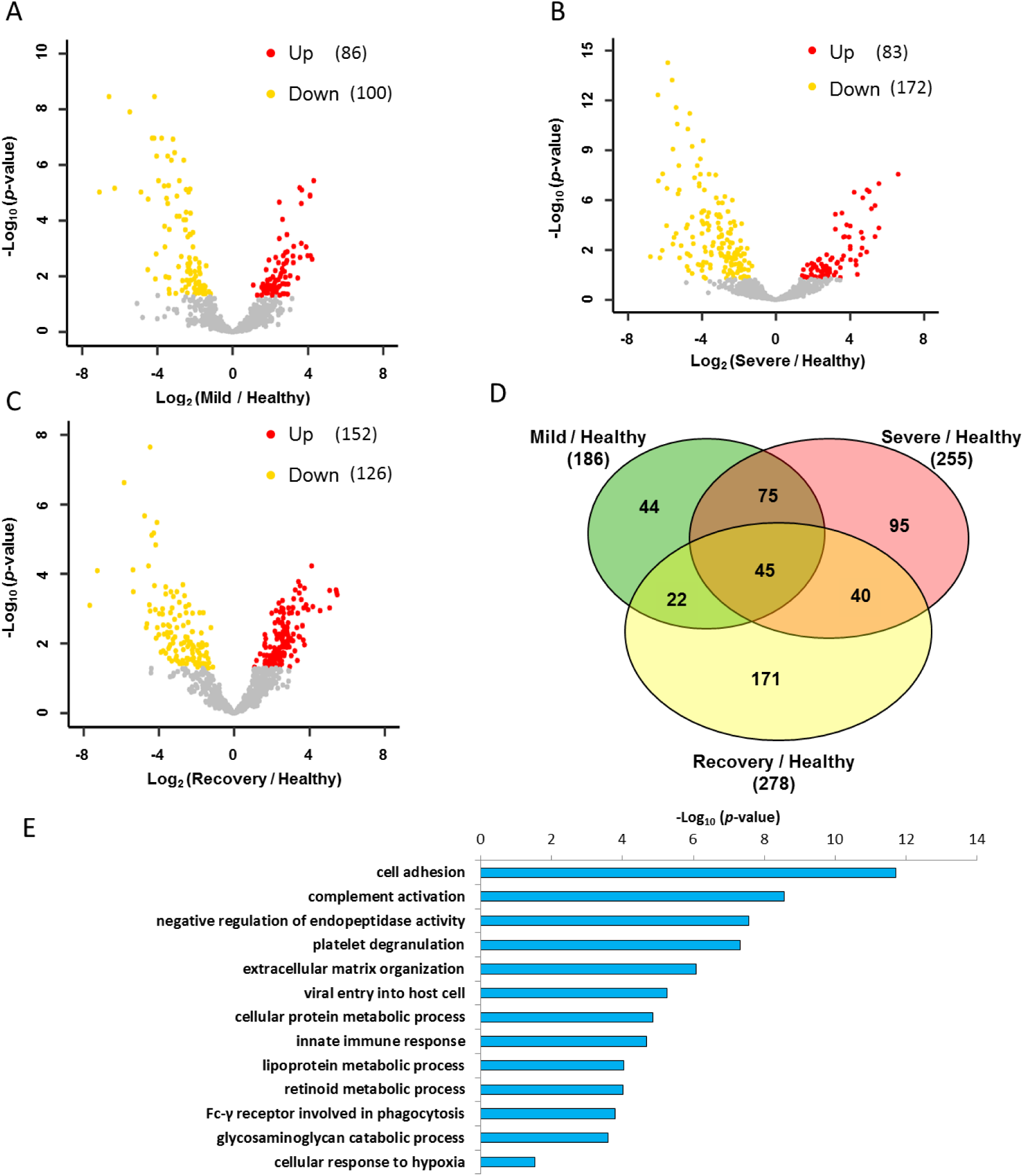
Function distribution of dysregulated proteins in COVID-19 patients. (A&B&C) The volcano plots of the up-regulated and down-regulated proteins in different groups. Proteins with *p*-Value lower than 0.05 and fold change ≥ 2 were considered as significantly differential expression. (D) Venn diagrams of differential proteins in mild, severe COVID-19 patients and recovery patients compared with healthy volunteers. (E) The GO analysis of dysregulated proteins in the COVID-19 patients.

To identify specific proteins to distinguish the mild from severe type of COVID-19 patients, we clustered the commonly identified proteins for all of these four datasets into 16 significant discrete clusters with the quantified values (Figure S7) through mFuzz [24]. We chose the cluster 2 and 11 as severe COVID-19 up-changed from mild COVID-19 (Figure 5A). Combined the filter results and significantly changed proteins from figure 4D, we identified 56 unique proteins conforming to the criteria. These proteins were highly associated with the complement activation, regulation of immune response, cellular oxidant detoxification, cellular response to hypoxia and oxidative stress-induced apoptosis, which might reflect the pathogenesis of the severe COVID-19. These results are consistent with the recent reported sera proteomics [20]. We also chose the cluster 1 and 12 as the down-regulated filter of the severe COVID-19 from mild COVID-19 as well (Figure 5C). These filtered proteins were highly associated with the platelet degranulation, glucose metabolic process, protein metabolic process and lipid metabolic and transport pathways.

**Figure 5.**
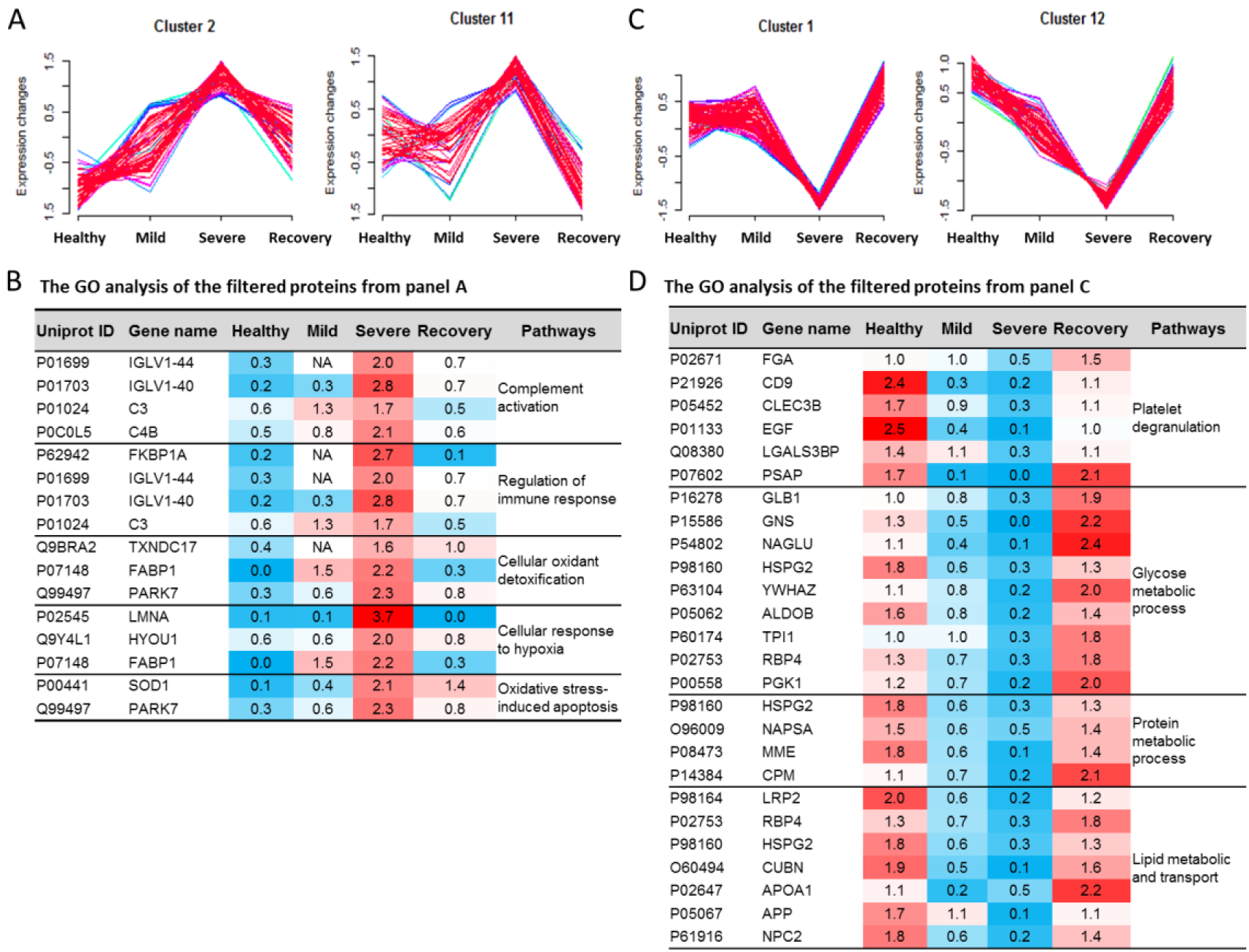
Clustering of commonly identified proteins illustrated specific clusters of proteins in COVID-19 patients. The numbers 1-4 stands for the Health, Mild, Severe and Recovery, respectively. (A) The cluster 2 and 11 stands for the up-regulated trends uniquely in the severe type of COVID-19. (B) The GO analysis of the filtered proteins from panel A. (C) The cluster 1 and 12 stands for the down-regulated trends uniquely in the severe type of COVID-19. (D) The GO analysis of the filtered proteins from panel C.

The molecular features used to distinguish the patient type (M and S) in our classifier (Figure 5B and 5D, Tables S4-5) contain several potential biomarkers which were highly associated with the clinical characteristics of mild and severe COVID-19. For example, the hypoxia up-regulated protein 1 (HYOU1) belonging to cluster 2 was more than three-fold higher in the severe COVID-19 (Figure 5B). HYOU1 plays a pivotal role in cyto-protective cellular mechanisms triggered by oxygen deprivation and is highly expressed in tissues such as liver and pancreas that contain well-developed endoplasmic reticulum and also regulates large amounts of secretory proteins [25, 26]. Patients with hypoxia warrant more attention to their intravascular coagulation, such as the elevated levels of D-dimer, a blood marker of excess clotting. It was reported that the heparin could boost patients’ low oxygen levels regardless of whether they were struggling to breathe [27]. In this study, we found that the heparin cofactor 2 (SERPIND1) belonging to cluster 10 was specially up-regulated more than four-fold higher in the mild and two-fold higher in the severe COVID-19 (Table S4). SERPIND1, also known as heparin cofactor II, is a glycoprotein in human plasma that inhibits thrombin and chymotrypsin, and the rate of inhibition of thrombin is rapidly increased by Dermatan sulfate (DS), heparin (H) and glycosaminoglycans (GAG) [28, 29]. We speculated that the SERPIND1 could be the protective response to reduce the risk of excess intravascular coagulation in the COVID-10 patients.

We also found that the cyclic AMP-responsive element-binding protein 3-like protein 3 (CREB3L3) belonging to cluster 10 (Figure S7) was specially up-regulated in the M-COVID (Table S4). In acute inflammatory response, CREB3L3 may activate expression of acute phase response (APR) genes, which was activated in response to cAMP stimulation [30]. This might be the protective mechanism for body to fight against the virus.

For the down-regulated molecular clusters, the proteins related with platelet degranulation was also reported in the sera proteomics recently [20]. Additionally, the down-regulated pathway of lipid metabolic and transport in the COVID-19 patients caused our attention. The cholesterol homeostasis was reported to impact COVID-19 prognosis, virus entry and the antiviral therapies [31]. In our data, the lipid metabolism and transporting, including the cholesterol homeostasis, were down-regulated in the S-COVID (Figure 5D and Table S5). The proteins NPC intracellular cholesterol transporter 2 (NPC2), apolipoproteins A1 (APOA1), and Cubilin (CUBN) were changed with the similar trends (Figure 5D). These results indicated that after the SARS-CoV-2 infection, the lipoprotein-mediated cholesterol uptake and transporting was disordered. Our study suggests that more characteristic molecular changes at protein levels can be used to build a predictive filter for the prospective identification of severe cases and shed light on the understanding of COVID-19 pathophysiology.

## DISCUSSION

The COVID-19 pandemic caused by SARS-CoV-2 is not only putting huge pressure on global healthcare, but also having a devastating impact on the economy and society. Although much effort towards COVID-19 diagnostics and treatment has been made, the mortality of this infectious disease has not been significantly improved because of the limited mechanistic understanding of the pathogenesis [2, 32]. Patients progress into the S-COVID often face very limited treatment options [8, 9]. Imaging technology, such as CT has been widely used to diagnose the COVID-19 patients, but suffers from high cost and demand for technical expertise, which is only accessible to those who live in developed countries. There is an urgent need for low-cost and reliable diagnostic techniques to estimate and predict the transition of severe COVID-19 patients from mild COVID-19 ones.

Urine is one of the most frequently studied biomaterials for biomarkers of human diseases in proteomics study because of its accessibility. It is less complex and has a relatively lower dynamic range with less technical challenges compared to blood [33–36]. Recently, it was demonstrated to effectively predict lung cancer [21]. Here, we assayed human urine samples from 32 healthy donors, three M-COVID, three S-COVID patients and two COVID-19 recovery person. Our study demonstrated that urine profiling could separate the healthy control from COVID-19 patients and also tell recovery person from COVID-19. Specific proteome features for M-COVID and S-COVID patients were detected in the urine samples. This is the first study to establish a link between the urine proteome and the understanding of the COVID-19 pathophysiology. Though the number of samples collected in this study was small, the obtained findings are consistent with the previous blood study [20], which supports the accuracy of our results. Our new findings would have potential implications for clinical diagnosis and treatment. We concluded that urine proteome is an important source that warrants more attentions for the understanding of COVID-19.

Given the possibility of using proteome as a diagnostic tool we have shown, we are also aware of several limitations in our COVID-19 urine proteomics study. First, only limited number of COVID-19 patient samples were included in our proteomics study, future study to include more samples will likely mitigate the possible sampling bias. Second, the current COVID-19 patients were only categorized as M-COVID and S-COVID. We could not obtain intermediate type of COVID-19 patient samples to improve the resolution to predict the trend and progress of atypical pneumonia caused by SARS-CoV-2 due to the limited number of patients. Third, the batch effect of the sample processing and proteomics analysis may cause some deviations. Some healthy control datasets were generated before, though with the similar experiment processes. We recommended that the urine proteomics researches of the healthy, COVID-19 and their corresponding recovery samples were performed meanwhile if possible. Fourth, patients were subjected to different antiviral drug treatments, compounded with their age, preexisting health conditions as well as wide range of days of onset symptoms, it might cause some bias to the conclusion at this stage.

Altogether, our data demonstrate that a urine proteome-based proteomics study can reliably and sensitively differentiate COVID-19 patients from healthy people. It might be able to serve as a powerful tool to help scientists and clinicians fight the COVID-19 pandemic.

## MATERIALS AND METHODS

### Urine samples

All COVID-19 patients were diagnosed according to the Diagnosis and management plan of pneumonia with new coronavirus infection (Trial Version 6) in the Fifth Medical Center of Chinese PLA General Hospital, Beijing, China, between February 18 and March 3, 2020. According to the Diagnosis standards, those patients were classified as clinically severe type infection empirically based on a set of clinical characteristics, such as dyspnea, respiratory rate (RR ≥ 30 times/min), mean oxygen saturation (≤ 93%, resting state) or arterial blood oxygen partial pressure/oxygen concentration (PaO_2_/FiO_2_ ≤ 300 mmHg), and/or lung infiltrates > 50% within 24-48 hours. The patients classified as mild type infection were mainly manifested with the symptoms of fever, non-pneumonia or mild pneumonia cases. A total of 7 urine specimens from COVID-19 patients were collected. One of them was discarded because of severe renal failure. Patients with underlying diseases except renal dysfunction are indicated in Figure 1A. The patients are aged from 59 to 78 years old (Figure 1A). Among the six analyzed samples, two of them are female.

A total of 32 urine specimens from healthy controls (CTL, n=32) were collected at Beijing Proteome Research Center, Beijing, China. The midstream of the morning urine was obtained for this study. Healthy controls are aged from 22 to 39 years old without any underlying disease. Among them, 11 are female (Figure S1).

All participants have provided signed informed consent and samples were collected with ethics approval from institutional review board (IRB) from the Fifth Medical Center of Chinese PLA General Hospital and Beijing Proteome Research Center. Our research strictly followed the standards indicated by the Declaration of Helsinki.

### Proteomics sample preparation and LC-MS/MS analysis

Human urine proteomics samples were prepared as described previously with slight modification [21, 33–36]. Briefly, 1 mL urine samples were centrifuged at 2,000 g for 4 min to remove cell debris before reduced with 5 mM dithiotheitol (DTT) at 56°C for 30 min, which could also inactivate the virus. The treated samples were alkylated with 10 mM iodoacetamide in dark at room temperature for 30 min. The supernatant was loaded into a 10 kDa ultrafiltration tube and the larger molecular weight proteins (proteome) were separated from the endogenous peptides (peptidome) by centrifugation. Proteome samples were digested with trypsin at 37 °C for 14h then the digestion reaction was terminated by 1% formic acid (FA). The digested peptides were desalted through a StageTip [37, 38] and dried before LC-MS/MS analysis.

The dried peptides were dissolved with 20 μL loading buffer (1% formic acid, FA; 1 % acetonitrile, ACN). 6 μL sample was taken for LC-MS/MS analysis on an Orbitrap Fusion Lumos coupled with EASY-nLC 1200 (Thermo Fisher Scientific, Waltham, MA, USA).

The samples were loaded onto a self-packed trap column (2 cm × 150 μm) and then separated by a capillary column (15 cm × 150 μm), both packed with C18 reverse phase particle (1.9 μm, Phenomenex, Torrance, California, USA). The peptides were eluted with a 120 min nonlinear gradient: 6% B for 10 min, 9-14% B for 15 min, 14-30% B for 50 min, 30-40% B for 30 min, 40-95% B for 3 min, 95% B for 7 min, 95-6% B for 1 min, 6% B for 4 min (Buffer A, 0.1% FA in ddH_2_O; Buffer B, 0.1% FA and 80% ACN in ddH_2_O; flow rate, ~600 nL/min).

The parameters for MS detecting were as follows: The full MS survey scans were performed in the ultra-high-field Orbitrap analyzer at a resolution of 120,000 and trap size of 500,000 ions over a mass range from 300 to 1,400 m/z. MS/MS scan were detected in IonTrap and the 20 most intense peptide ions with charge states 2 to 7 were subjected to fragmentation via higher energy collision-induced dissociation (1×10^4^ AGC target, 35 ms maximum ion time).

### Data processing and label-free quantification

The raw files were searched with MaxQuant software (v1.5.3.0) against the database composed of Human fasta downloaded from Swiss-Prot (version released in 2020.02) and the SARS-CoV-2 virus fasta downloaded from NCBI (RefSeq GCF_009858895.2). Mass tolerance of the first search for precursor ions was set to 20 ppm. Full cleavage by trypsin was set and a maximum of two missed cleavages was allowed. The protein identification must met the following criteria: (1) the peptide length≥7 amino acids; (2) the FDR≤1% at the PSM, peptide and protein levels.

The peptides were quantified by the peak area derived from their MS1 intensity with MaxQuant software [39]. The intensity of unique and razor peptides was used to calculate the protein intensity. The intensity based absolute quantification (iBAQ) algorithm was used as protein quantification value [40]. In order to exclude the influence of differences in sample sizes and loading amounts for MS analysis, we used median value of each sample to normalize protein iBAQ values [41]. All missing values were substituted with the minimal value.

### Statistical analyses

Overlapped 1008 proteins were used for the subsequent statistical analysis. Pearson correlation analysis of all datasets was realized by Perseus [42]. Differential proteins were filtered using R package limma (version 3.34.9). The significantly differentially expressed proteins were selected using the criteria of adjusted p value less than 0.05 and log_2_ FC larger than 1. Proteins were clustered using R package mFuzz (version 2.46.0) into 16 significant discrete clusters.

### Pathway analyses

The function of differential proteins was analyzed in David Bioinformatics Resources (https://david.ncifcrf.gov/) and Human Protein Atlas (http://www.proteinatlas.org/) platforms including tissue-specific enrichment, molecular function, biological process, cellular component, etc.

## Data Availability

The accession numbers for the mass spectrometry proteomics data reported in this paper are the iProX (https://www.iprox.org/) dataset identifier: IPX0002166000. All the data will be publicly released upon publication.

## DATA AVAILABILITY

The mass spectrometry proteomics data have been deposited to the ProteomeXchange Consortium (http://proteomecentral.proteomexchange.org) via the iProX partner repository[43]. The accession numbers for the mass spectrometry proteomics data reported in this paper are the iProX (https://www.iprox.org/) dataset identifier: IPX0002166000. All the data will be publicly released upon publication.

## ACKNOWLEDGEMENTS

This study was funded by the MOST (2017YFC0906600 & 2017YFA0505002), the National Natural Science Foundation of China (31700723, 31670834, 31870824, 91839302 & 31901037), the Innovation Foundation of Medicine (16CXZ027, BWS17J032, AWS17J008 & 19SWAQ17), National Megaprojects for Key Infectious Diseases (2018ZX10302302), the Foundation of State Key Lab of Proteomics (SKLP-K201704), and the grant for Research Unit of Proteomics & Research and Development of New Drug of Chinese Academy of Medical Sciences (2019RU006).

## AUTHOR CONTRIBUTIONS

P.X., C.B., and H.W. designed the experiment and supervised this project. Y.L., Y.W. and W.S. performed the proteomic experiments and data analysis with the help of Y.Z., P.C., and L.C.. H.L. and C.B. collected the samples and clinical data with the help of L.Z., Z.L. and N.L.. Data were interpreted and presented by all co-authors. P.X., Y.L., B.D., and Y.W. wrote the manuscript with inputs from co-authors.

